# Disparities in deworming coverage between children with and without disabilities: insights from the DeWorm3 trial in India

**DOI:** 10.1101/2025.06.05.25329067

**Authors:** Shanquan Chen, Rohan Michael Ramesh, Kumudha Aruldas, Saravanakumar Puthupalayam Kaliappan, Bobeena Rachel Chandy, Beena Koshy, Smitha Jasper, Katherine E. Halliday, Judd L. Walson, William Oswald, Sitara Swarna Rao Ajjampur, Hannah Kuper

**Affiliations:** International Centre for Evidence in Disability, London School of Hygiene & Tropical Medicine, London, United Kingdom, WC1E 7HT; The Wellcome Trust Research Laboratory, Division of Gastrointestinal Sciences, Christian Medical College, Vellore, India; Department of Physical Medicine and Rehabilitation, Christian Medical College, Tamil Nadu, India; Department of Developmental Paediatrics, Christian Medical College, Tamil Nadu, India; Department of Ophthalmology, Christian Medical College, Tamil Nadu, India; Faculty of Infectious and Tropical Diseases, London School of Hygiene & Tropical Medicine, London, United Kingdom; Departments of International Health, Medicine and Pediatrics, Johns Hopkins Bloomberg School of Public Health, Baltimore, USA, MD 21205; RTI International

**Keywords:** mass drug administration, deworming, disability, children, community-based program, school-based program, India

## Abstract

**Background:** Children with disabilities experience lower school enrollment rates and may be systematically excluded from school-based deworming programs. The DeWorm3 trial provided an opportunity to evaluate whether community-wide mass drug administration (cMDA) could reduce these disparities by reaching children outside the education system.

**Methods:** We conducted a secondary analysis of the data from the DeWorm3 trial in Tamil Nadu, India. Children aged 5-17 years (n=82,417) who participated in at least one of the six rounds of cMDA were included. Disability was assessed using the UNICEF/Washington Group’s Child Functioning Module. Mixed-effects logistic regression models examined associations between disability status and treatment coverage, adjusting for sociodemographic factors.

**Results:** Children with disabilities (1.1% of the population) had higher unadjusted odds of not receiving deworming treatment compared to children without disabilities (OR=1.49, 95% CI: 1.19-1.84), though this association was attenuated after adjustment (adjusted OR=1.10, 95% CI: 0.86-1.37). Among school-attending children who received treatment, those with disabilities had significantly lower odds of receiving school-based treatment (adjusted OR=0.57, 95% CI: 0.44-0.73). Subgroup analyses revealed that age, school attendance, parental marital status, geographic location, and the COVID-19 pandemic significantly modified these associations.

**Conclusion:** While community-wide mass drug administration reduced some disparities in deworming coverage, children with disabilities remained less likely to receive school-based treatment. Public health programs should combine school-based approaches with targeted community outreach strategies to ensure equitable inclusion of children with disabilities, particularly those experiencing multiple vulnerabilities.

## Introduction

Globally, an estimated 200 million children aged 5-17 years live with disabilities[1]. These children frequently face exclusions across multiple dimensions, including education and access to health interventions[2, 3]. Soil-transmitted helminths (STH) remain a major public health concern, particularly in low- and middle-income countries, where over 1.5 billion people are affected[4]. Targeted deworming programs are the cornerstone of STH control efforts aiming to reduce infection intensity and associated morbidity in risk groups through preventive chemotherapy[4, 5]. However, traditional school-based deworming (SBD) programs, such as India’s National Deworming Day (NDD), primarily target pre-school and school aged children attending school, potentially excluding children with disabilities—many of whom are not enrolled in anganwadis (informal state-run learning centers for preschool children established under the Integrated Child Development Scheme) and school. Such systematic exclusions not only reinforce health inequities but may also hinder the overall success of elimination efforts.

The DeWorm3 trial represents a paradigm shift from STH control to evaluating the feasibility of STH transmission interruption through community-wide mass drug administration (cMDA) compared with school-based deworming [6, 7]. cMDA aims to treat all individuals within a community, including children aged >1 year, regardless of school enrollment, thereby potentially addressing the limitations of school-based approaches. However, the effectiveness of cMDA in reaching vulnerable populations such as children with disabilities remains underexplored. This is an important topic as children with disabilities are disproportionately impacted by health disparities due to barriers in accessing care, heightened stigma, and misconceptions regarding their health needs[8–10]. The integration of disability-inclusive approaches within MDA campaigns is critical for achieving equitable health outcomes and sustaining transmission interruption goals[4]. The paucity of evidence on deworming coverage among children with disabilities, particularly in community-based and school-based settings, underscores the need for targeted research.

This study aims to assess disparities in deworming treatment coverage between children with and without disabilities within the context of the DeWorm3 trial in southern India. Specifically, the study seeks to answer the following research questions: first, what is the likelihood of children with disabilities receiving deworming treatment compared to children without disabilities during cMDA? Second, among school-attending children who received deworming, what is the likelihood of children with disabilities being treated at school rather than in the community compared with children without disabilities? By addressing these questions, this study seeks to inform the design and implementation of equitable deworming strategies that include all children, particularly those with disabilities.

## Methods

### Study Design and Participants

This study is a secondary analysis of data from the DeWorm3 trial, a multi-country cluster-randomized community-based study designed to evaluate the feasibility of interrupting soil-transmitted helminths (STH) transmission through cMDA. Details of the trial design, methods, and results have been described extensively elsewhere[6, 7].

The analysis focuses on intervention clusters in India, comprising 40 clusters in Tamil Nadu’s Timiri and Jawadhu Hills blocks[5, 11–14]. These clusters included a total population of approximately 141,000, residing in 36,536 households across 219 villages in Timiri and 154 villages in Jawadhu Hills[5, 11–14]. This data represents all 40 clusters in the study area. The residents in this geographical area are characterized by predominantly rural (77%) and tribal (23%) residents, with limited sanitation access (34.6%) and significant engagement in agricultural activities (20% in Timiri, 90% in Jawadhu Hills) [5, 11–14].

Eligible participants for this analysis were children aged 5–17 years who had participated in at least one round of cMDA.

### Data Collection

Data were collected as part of the routine activities of the DeWorm3 trial by adhering to rigorous and systematic protocols. The baseline census, conducted between 2017 and 2018, enumerated all households and individuals within the intervention clusters[5, 15]. This census captured extensive demographic, socioeconomic, and geographic data, including age, gender, disability status, education levels, and household wealth indicators. Each household received a unique identification card with a barcode to enable precise tracking in subsequent data collection rounds. Annual updates to the census were undertaken to account for population changes such as births, deaths, and migration, ensuring accurate denominators for coverage and treatment analyses.

During the six rounds of cMDA conducted for three years (2018 to 2020), albendazole was provided to eligible individuals aged 1–99 years. For children, deworming treatment was initially offered through the National Deworming Day (NDD) program administered by teachers at schools. Following NDD activities, community drug distributors (CDDs) conducted cMDA, which targeted individuals not treated during NDD (identifiable by the absence of ink marks on fingers). Trained community drug distributors (CDDs), supported by study officers, delivered the drug directly to households. Treatment administration was directly observed and electronically recorded using a custom-designed treatment register. Efforts were made to maximize treatment coverage through repeated household visits for individuals initially unavailable. If absence persisted, treatment tablets were left with the head of the household, and acceptance was documented. For school-attending children, deworming during India’s National Deworming Day (NDD) program was verified by inspecting ink marks applied on their fingers during school-based deworming sessions.

The DeWorm3 trial received ethical approval from the Institutional Review Boards (IRBs) of Christian Medical College (CMC), Vellore (10392 [INTERVEN]), the Human Subjects Division at the University of Washington (STUDY00000180), and the London School of Hygiene and Tropical Medicine Observational/Interventions Research Ethics Committee (12013). This trial was registered at ClinicalTrials.gov (NCT03014167). The consent procedures followed ethical guidelines appropriate to the context. Informed consent for participation in the trial, including census and drug administration activities, was obtained from the head of each household. Written consent was collected from literate participants, while oral consent was documented via a thumbprint in the presence of a witness for those unable to write. Individual-level consent for specific activities beyond household participation was deemed unnecessary for this study, as data were derived from routine trial operations.

### Outcomes and Measurement

The primary outcomes of this study were: (1) the disability-related disparity in treatment coverage during community-wide mass drug administration (cMDA), measured as the likelihood of children with disabilities receiving treatment compared to children without disabilities, as recorded in cMDA treatment registers; and (2) the disparity in school-based treatment among dewormed, school-attending children, evaluated as the likelihood of children with disabilities receiving treatment at school compared to their typically developing peers. The latter outcome was assessed through self-reported data collected during surveys conducted after the first four cMDA rounds, as school-based deworming was not implemented during the final two rounds due to the COVID-19 lockdown. During the second COVID-19 round, National Deworming Day (NDD) activities were conducted in community settings rather than schools.

Disability status was assessed using the Child Functioning Module (CFM), a tool validated for use in surveys with mothers or primary caregivers as proxy respondents[16]. Developed by UNICEF in collaboration with the Washington Group on Disability Statistics, CFM is grounded in the World Health Organization’s International Classification of Functioning and the biopsychosocial model of disability[17]. The module has undergone extensive expert review and testing across multiple countries to ensure the quality of its questions and their cultural appropriateness for respondents with varying linguistic backgrounds and experiences of disability[17]. For children aged 5–17 years, the CFM evaluates functional difficulties across several domains, including vision/seeing, hearing, mobility/walking, self-care, communication/comprehension, learning, remembering, focusing attention/concentrating, coping with change, controlling behaviour, and relationships/making friends. Details of the module’s structure and administration are described elsewhere[18]. Functional difficulties in each domain were measured using a four-point response scale: “no difficulty,” “some difficulty,” “a lot of difficulty,” or “cannot do at all.” Following UNICEF’s definition, children were classified as having a functional disability if they were reported to have “a lot of difficulty” or “cannot do at all” in at least one domain[16].

### Covariates

The following variables were considered as covariates: individual-level sociodemographic factors, such as age, gender, and school attendance, as well as household-level characteristics, including the highest education level attained within the family, parental marital status, and household size (≥5 members). Additional factors considered were caste, rural residence (yes/no), study site, and survey round (1–6), to account for geographic and temporal variations in the analysis.

### Statistical Analysis

Categorical variables were reported as frequencies and percentages, while continuous variables were reported as means with standard deviations (SD). Differences in outcomes were evaluated using two-tailed t-tests for continuous variables and chi-square tests for categorical variables.

A mixed-effects logistic regression model was employed to investigate the association between disability status and likelihood of not receiving deworming treatment. The dependent variable was untreated status (yes or no), with disability status (yes or no) as the primary independent variable. The model was adjusted for sociodemographic covariates, including age, sex, school attendance, household educational attainment, parental marital status, household size, socioeconomic status, caste, residential location, study site, and survey round. Random intercepts were included for participants’ unique IDs to account for within-individual clustering. Results are reported as odds ratios (OR) with 95% confidence intervals (CI). Subgroup analyses were conducted by repeating the regression model stratified by covariates, and the interaction effects between disability status and subgroup variables were assessed using the interaction terms within the overall model.

The analysis was repeated to examine the association between disability status and the likelihood of receiving school-based deworming treatment among children who attended school and had received deworming.

Missing data were predominantly observed in variables such as age (0.23%), gender (0.01%), school attendance (0.24%), religion (1.81%), caste (2.20%), socioeconomic status (<0.01%), household education (0.03%), parental marital status (<0.01%), and household size (0.23%). To minimize bias due to missing data, multiple imputations using chained equations with 5mputations were performed.

All statistical analyses were conducted using R software (version 4.3.0), with statistical significance set at P<0.05.

The Euclidean distance to schools the children attended was analyzed using ArcGIS Pro (version 3.3.2)

### Patient and Public Involvement

This study is a secondary analysis of data collected during the DeWorm3 trial. While patients and the public were not involved in the design, analysis, or dissemination of this specific secondary analysis, the broader DeWorm3 trial incorporated community engagement as a core component. Community leaders, parents, and local health stakeholders were actively engaged during trial planning and implementation to ensure culturally appropriate delivery and acceptability of mass drug administration activities. Community drug distributors, drawn from the local population, played a central role in treatment delivery and community sensitisation. The study team also conducted formative qualitative research to understand local perspectives and incorporated these findings into the implementation strategy. Although children with disabilities and their caregivers were not specifically involved in shaping this analysis, our findings highlight the importance of future studies incorporating participatory approaches to address equity in neglected tropical disease programmes.

## Results

**Table 1** presents the characteristics of 82,417 children aged 5-17 years from the DeWorm3 trial conducted in India. Among these children, 58.8% were aged 12-17 years, 47.8% were female, and 89.9% had attended school. Most children (93.2%) received deworming treatment, with 48.4% being treated in school and 51.6% in the community. Children with disabilities constituted 1.1% of the total population, with varying types of functional difficulties, including seeing (0.1%), hearing (0.1%), mobility/walking (0.4%), self-care (0.4%), communication (0.5%), and others. The majority of children came from households where parents were married (90.3%), lived in rural areas (85.2%), and belonged to the Hindu religion (96.3%). The study population was primarily from the Timiri site (71.9%) compared to Jawadhu Hills (28.1%). During the COVID-19 pandemic period (rounds 5-6), there was a notable shift from school-based to community-based treatment delivery, with virtually no school-based treatments during these rounds. The basic description by outcomes was also presented in **Table 1**.

**Table 1.** Basic characteristics, by deworm treatment status and by treated place. Data was reported as the mean (standard deviation) or number (percentage). P-values were derived using two-tailed tests for continuous variables and two-tailed chi-square tests for categorical variables. For categorical variables with expected cell counts less than 5, Fisher’s exact test was employed.

|  | All<br>(n = 82,417) | Not treated<br>(n = 5,625) | Treated<br>(n = 76,792) | p value | Treated in school<br>(n = 38,430) | Treated in community<br>(n = 38,362) | p value |
| --- | --- | --- | --- | --- | --- | --- | --- |
| **Age** |  |  |  |  |  |  |  |
| 5-11 years | 33959 (41.2%) | 2070 (36.8%) | 31889 (41.5%) | <0.01 | 16411 (42.7%) | 15478 (40.3%) | <0.01 |
| 12-17 years | 48458 (58.8%) | 3555 (63.2%) | 44903 (58.5%) |  | 22019 (57.3%) | 22884 (59.7%) |  |
| **Sex (= female)** | 39411 (47.8%) | 2578 (45.8%) | 36833 (48.0%) | <0.01 | 19198 (50.0%) | 17635 (46.0%) | <0.01 |
| **Attending school (= yes)** | 74123 (89.9%) | 4165 (74.0%) | 69958 (91.1%) | <0.01 | 37153 (96.7%) | 32805 (85.5%) | <0.01 |
| **Highest education degree of family members** |  |  |  |  |  |  |  |
| No education | 19515 (23.7%) | 2340 (41.6%) | 17175 (22.4%) | <0.01 | 8057 (21.0%) | 9118 (23.8%) | <0.01 |
| Any primary | 15241 (18.5%) | 904 (16.1%) | 14337 (18.7%) |  | 7252 (18.9%) | 7085 (18.5%) |  |
| Any middle | 17589 (21.3%) | 881 (15.7%) | 16708 (21.8%) |  | 8711 (22.7%) | 7997 (20.8%) |  |
| Any secondary or higher | 30072 (36.5%) | 1500 (26.7%) | 28572 (37.2%) |  | 14410 (37.5%) | 14162 (36.9%) |  |
| **Parents marital status** |  |  |  |  |  |  |  |
| Married | 74447 (90.3%) | 4865 (86.5%) | 69582 (90.6%) | <0.01 | 34971 (91.0%) | 34611 (90.2%) | <0.01 |
| Previously or never married | 7970 (9.7%) | 760 (13.5%) | 7210 (9.4%) |  | 3459 (9.0%) | 3751 (9.8%) |  |
| **Household size (>=5)** | 49703 (60.3%) | 3550 (63.1%) | 46153 (60.1%) | <0.01 | 23396 (60.9%) | 22757 (59.3%) | <0.01 |
| **Member in the family temporarily migrated (=yes)** | 504 (0.6%) | 496 (8.8%) | 8 (0.0%) | <0.01 | 6 (0.0%) | 2 (0.0%) | 0.29 |
| **Distance to school (<= 1 km)** |  |  | 34792 (49.7%) |  | 18544 (49.9%) | 16248 (49.5%) | 0.31 |
| **Household socioeconomic status** |  |  |  |  |  |  |  |
| Poor (Lower 3 quintiles) | 49993 (60.7%) | 2295 (40.8%) | 47698 (62.1%) | <0.01 | 24057 (62.6%) | 23641 (61.6%) | 0.01 |
| Rich (Upper 2 quintiles) | 32424 (39.3%) | 3330 (59.2%) | 29094 (37.9%) |  | 14373 (37.4%) | 14721 (38.4%) |  |
| **Religion (= Hinduism (India))** | 79367 (96.3%) | 5407 (96.1%) | 73960 (96.3%) | 0.49 | 37142 (96.6%) | 36818 (96.0%) | <0.01 |
| **Caste** |  |  |  |  |  |  |  |
| Most backward caste | 19279 (23.4%) | 884 (15.7%) | 18395 (24.0%) | <0.01 | 9596 (25.0%) | 8799 (22.9%) | <0.01 |
| Scheduled tribes | 22362 (27.1%) | 2848 (50.6%) | 19514 (25.4%) |  | 9372 (24.4%) | 10142 (26.4%) |  |
| Scheduled caste | 20510 (24.9%) | 909 (16.2%) | 19601 (25.5%) |  | 9863 (25.7%) | 9738 (25.4%) |  |
| Backward caste | 19534 (23.7%) | 950 (16.9%) | 18584 (24.2%) |  | 9266 (24.1%) | 9318 (24.3%) |  |
| Higher caste | 732 (0.9%) | 34 (0.6%) | 698 (0.9%) |  | 333 (0.9%) | 365 (1.0%) |  |
| **Residence place (= rural)** | 70251 (85.2%) | 4891 (87.0%) | 65360 (85.1%) | <0.01 | 32938 (85.7%) | 32422 (84.5%) | <0.01 |
| **Site** |  |  |  |  |  |  |  |
| Timiri | 59286 (71.9%) | 2739 (48.7%) | 56547 (73.6%) | <0.01 | 28692 (74.7%) | 27855 (72.6%) | <0.01 |
| Jawadhu Hills | 23131 (28.1%) | 2886 (51.3%) | 20245 (26.4%) |  | 9738 (25.3%) | 10507 (27.4%) |  |
| **Disabled (= Yes)** | 940 (1.1%) | 92 (1.6%) | 848 (1.1%) | <0.01 | 264 (0.7%) | 584 (1.5%) | <0.01 |
| Seeing | 92 (0.1%) | 11 (0.2%) | 81 (0.1%) | 0.08 | 30 (0.1%) | 51 (0.1%) | 0.03 |
| Hearing | 106 (0.1%) | 20 (0.4%) | 86 (0.1%) | <0.01 | 24 (0.1%) | 62 (0.2%) | <0.01 |
| Mobility/walking | 327 (0.4%) | 44 (0.8%) | 283 (0.4%) | <0.01 | 58 (0.2%) | 225 (0.6%) | <0.01 |
| Self-care | 329 (0.4%) | 43 (0.8%) | 286 (0.4%) | <0.01 | 45 (0.1%) | 241 (0.6%) | <0.01 |
| Communication | 381 (0.5%) | 54 (1.0%) | 327 (0.4%) | <0.01 | 69 (0.2%) | 258 (0.7%) | <0.01 |
| Learning | 321 (0.4%) | 43 (0.8%) | 278 (0.4%) | <0.01 | 56 (0.1%) | 222 (0.6%) | <0.01 |
| Remembering | 304 (0.4%) | 48 (0.9%) | 256 (0.3%) | <0.01 | 44 (0.1%) | 212 (0.6%) | <0.01 |
| Concentrating | 268 (0.3%) | 45 (0.8%) | 223 (0.3%) | <0.01 | 31 (0.1%) | 192 (0.5%) | <0.01 |
| Coping with change | 248 (0.3%) | 39 (0.7%) | 209 (0.3%) | <0.01 | 29 (0.1%) | 180 (0.5%) | <0.01 |
| Behaviour controlling | 295 (0.4%) | 42 (0.7%) | 253 (0.3%) | <0.01 | 47 (0.1%) | 206 (0.5%) | <0.01 |
| Making friends | 268 (0.3%) | 43 (0.8%) | 225 (0.3%) | <0.01 | 36 (0.1%) | 189 (0.5%) | <0.01 |
| **Survey round** |  |  |  |  |  |  |  |
| Round 1 | 11562 (14.0%) | 851 (15.1%) | 10711 (13.9%) | <0.01 | 7499 (19.5%) | 3212 (8.4%) | <0.01 |
| Round 2 | 12728 (15.4%) | 555 (9.9%) | 12173 (15.9%) | <0.01 | 9683 (25.2%) | 2490 (6.5%) | <0.01 |
| Round 3 | 13831 (16.8%) | 441 (7.8%) | 13390 (17.4%) | <0.01 | 10481 (27.3%) | 2909 (7.6%) | <0.01 |
| Round 4 | 14406 (17.5%) | 382 (6.8%) | 14024 (18.3%) | <0.01 | 10725 (27.9%) | 3299 (8.6%) | <0.01 |
| Round 5 | 14937 (18.1%) | 2240 (39.8%) | 12697 (16.5%) | <0.01 | 41 (0.1%) | 12656 (33.0%) | <0.01 |
| Round 6 | 14953 (18.1%) | 1156 (20.6%) | 13797 (18.0%) | <0.01 | 1 (0.0%) | 13796 (36.0%) | <0.01 |
| **COVID-19 pandemic (= yes)** | 29890 (36.3%) | 3396 (60.4%) | 26494 (34.5%) | <0.01 | 42 (0.1%) | 26452 (69.0%) | <0.01 |

**Figure 1** shows the treatment coverage patterns across the survey rounds for children with and without disabilities. Across all survey rounds, the proportion of children without disabilities who did not receive MDA treatment ranged from approximately 2.5% to 15.0%, whereas the corresponding proportion of children with disabilities ranged from approximately 5.0% to 13.0%. Panel A demonstrates that pre-COVID untreated proportions were low (<10%) for both groups, but higher for children with disabilities, followed by sharp increases in untreated children during the COVID-19 rounds (peaking at 25% vs. 15% in round 5). Panel B shows that among school-attending children who received treatment, those with disabilities were less likely to receive school-based treatment (60-70% vs 80-90%) during rounds 1-4 and instead were treated in the community, with treatment at schools ceasing for both groups during COVID-19 rounds 5-6.

**Figure 1.**
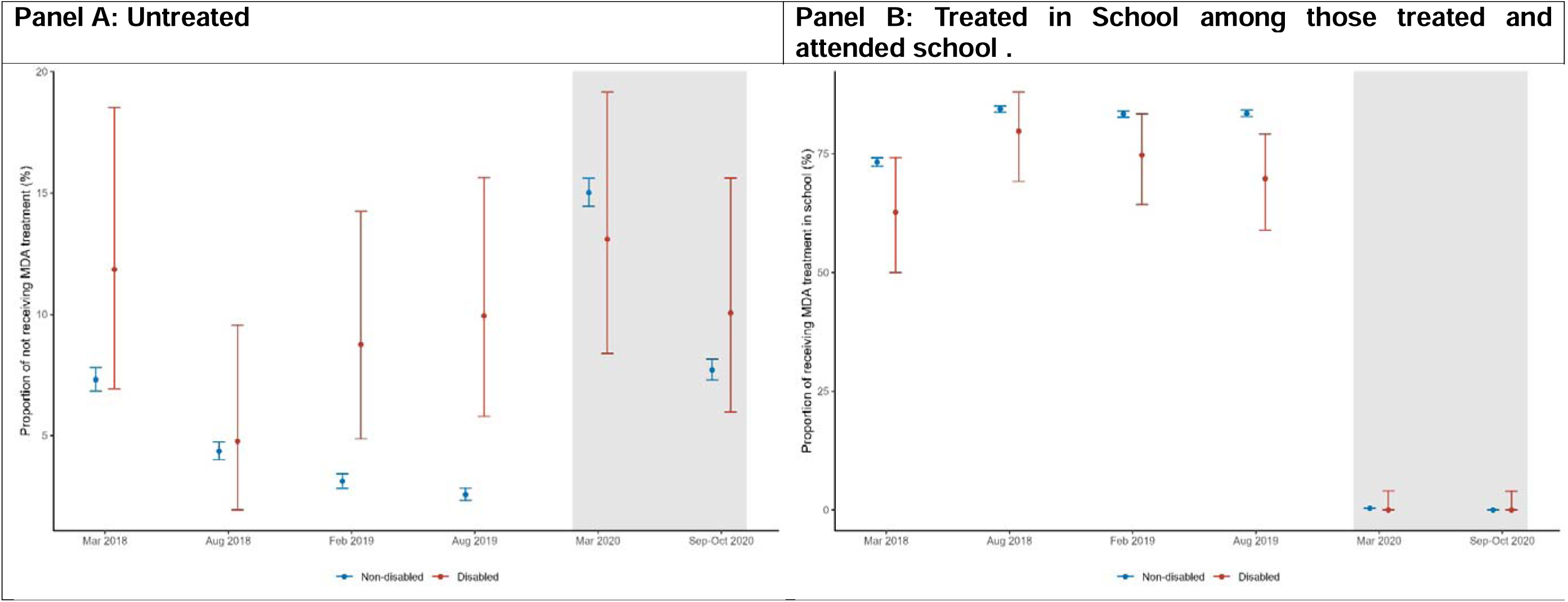
Proportion of untreated (Panel A) and treated in school (Panel B), by survey round and disability status. Points show the percentage, while vertical lines showing corresponding 95% confidence intervals. The shaded grey area denotes the period during which data were collected amid the COVID-19 pandemic.

**Table 2** presents the results of mixed-effects logistic regression analysis examining associations between disability status and deworming treatment non-receipt. While unadjusted analysis indicated children with disabilities had significantly higher odds of being untreated (OR=1.49, 95% CI: 1.19-1.84), this association was attenuated and became non-significant after adjusting for sociodemographic covariates including age, sex, school attendance, household educational attainment, parental marital status, household size, socioeconomic status, caste, residential location, site, and survey round (adjusted OR=1.10, 95% CI: 0.86-1.37). Subgroup analyses revealed significant interaction effects across several variables. Age demonstrated a significant interaction (interaction adjusted OR=2.05, 95% CI: 1.20-3.66), with older children (12-17 years) with disabilities showing disproportionately higher odds of non-treatment. School attendance also exhibited a significant interaction effect (interaction adjusted OR=1.66, 95% CI: 1.02-2.80), indicating that among school attendees, disability status was associated with increased odds of non-treatment. Parental marital status was another significant modifier (interaction adjusted OR=1.80, 95% CI: 1.00-3.15), with children with disabilities from previously or never married parents showing elevated odds of non-treatment. Geographic variation was evident through a significant site interaction (interaction adjusted OR=0.50, 95% CI: 0.31-0.81), suggesting differential treatment patterns between regions. The pandemic period demonstrated a significant interaction effect (interaction adjusted OR=0.39, 95% CI: 0.25-0.62), specifically showing that during the COVID-19 pandemic, the likelihood of non-treatment for children with disabilities was significantly reduced.

**Table 2.** Association between disability and non-receipt of deworming treatment, by overall and subgroups. A mixed-effects logistic regression analysis examining the association between disability status and non-receipt of deworming treatment . The model fitted with non-receipt of deworming treatment status (yes or no) as the dependent variable and disability status (yes or no) as the primary predictor, with adjustments for sociodemographic covariates including age, sex, school attendance, household educational attainment, parental marital status, household size, socioeconomic status, caste, residential location, site, and survey round. Random intercepts were included for participant’s unique ID. Results are presented as odds ratios (OR) with corresponding 95% confidence intervals (CI). Analyses include overall population estimates and stratified subgroup analyses. Interaction effects between disability status and subgroup variables were assessed by including the interactive term between disability status and corresponding subgroup variable. n/N refers to the number of untreated children (n) out of the total number of children in that category (N). For example, 92/940 indicates that 92 children with disability out of a total of 940 children with disability did not receive deworming treatment. p > 0.5, * p < 0.05, ** p < 0.01, *** p < 0.001

| Outcomes | Children with disability<br>n/N (%) | Children without disability<br>n/N (%) | Unadjusted OR | Adjusted OR | Interaction with subgroup |  |
| --- | --- | --- | --- | --- | --- | --- |
|  |  |  |  |  | Unadjusted OR | Adjusted OR |
| **Overall** | 92/940 (9.8%) | 5533/81477 (6.8%) | **1.49 [1.19, 1.84]\*\*\*** | 1.10 [0.86, 1.37] | - | - |
| **By age** |  |  |  |  |  |  |
| 5-11 years | 20/329 (6.1%) | 2050/33630 (6.1%) | 1.00 [0.61, 1.53] | 0.72 [0.43, 1.15] | Ref | Ref |
| 12-17 years | 72/611 (11.8%) | 3483/47847 (7.3%) | **1.70 [1.32, 2.17]\*\*\*** | **1.30 [0.99, 1.68]\*** | **1.71 [1.04, 2.93]\*** | **2.05 [1.20, 3.66]\*** |
| **By gender** |  |  |  |  |  |  |
| Females | 45/443 (10.2%) | 2533/38968 (6.5%) | **1.63 [1.18, 2.19]\*\*** | 1.14 [0.81, 1.58] | Ref | Ref |
| Males | 47/497 (9.5%) | 3000/42509 (7.1%) | **1.38 [1.00, 1.84]\*** | 1.02 [0.73, 1.40] | 0.85 [0.55, 1.31] | 0.86 [0.54, 1.36] |
| **Attended school** |  |  |  |  |  |  |
| No | 25/525 (4.8%) | 4140/73598 (5.6%) | 0.84 [0.55, 1.23] | 0.79 [0.50, 1.17] | Ref | Ref |
| Yes | 67/415 (16.1%) | 1393/7879 (17.7%) | 0.90 [0.68, 1.16] | 1.22 [0.90, 1.61] | 1.07 [0.67, 1.76] | **1.66 [1.02, 2.80]\*** |
| **Highest education degree of family members** |  |  |  |  |  |  |
| No education | 36/295 (12.2%) | 2304/19220 (12%) | 1.02 [0.71, 1.43] | 0.99 [0.67, 1.43] | Ref | Ref |
| Any primary | 24/212 (11.3%) | 880/15029 (5.9%) | **2.05 [1.30, 3.09]\*\*\*** | 1.35 [0.84, 2.10] | **2.01 [1.14, 3.49]\*** | 1.45 [0.80, 2.59] |
| Any middle | 8/150 (5.3%) | 873/17439 (5%) | 1.07 [0.48, 2.05] | 0.83 [0.36, 1.63] | 1.05 [0.44, 2.22] | 0.90 [0.37, 1.95] |
| Any secondary or higher | 24/283 (8.5%) | 1476/29789 (5%) | **1.78 [1.14, 2.65]\*\*** | 1.04 [0.64, 1.60] | **1.74 [1.00, 3.00]\*** | 1.18 [0.65, 2.09] |
| **Parents marital status** |  |  |  |  |  |  |
| Married | 72/815 (8.8%) | 4793/73632 (6.5%) | **1.39 [1.08, 1.76]\*\*** | 0.98 [0.75, 1.26] | Ref | Ref |
| Previously or never married | 20/125 (16%) | 740/7845 (9.4%) | **1.83 [1.10, 2.90]\*** | 1.65 [0.96, 2.72] | 1.31 [0.75, 2.22] | **1.80 [1.00, 3.15]\*** |
| **Household socioeconomic status** |  |  |  |  |  |  |
| Rich (Upper 2 quintiles) | 45/413 (10.9%) | 3285/32011 (10.3%) | 1.07 [0.77, 1.44] | 0.91 [0.64, 1.25] | Ref | Ref |
| Poor (Lower 3 quintiles) | 47/527 (8.9%) | 2248/49466 (4.5%) | **2.06 [1.50, 2.75]\*\*\*** | 1.25 [0.89, 1.71] | **0.52 [0.34, 0.80]\*\*** | 0.65 [0.41, 1.03] |
| **Residence place** |  |  |  |  |  |  |
| Rural | 10/100 (10%) | 724/12066 (6%) | 1.74 [0.85, 3.20] | 1.36 [0.64, 2.59] | Ref | Ref |
| Urban | 82/840 (9.8%) | 4809/69411 (6.9%) | **1.45 [1.15, 1.82]\*\*\*** | 1.08 [0.84, 1.37] | 1.20 [0.56, 2.30] | 1.08 [0.49, 2.14] |
| **Site** |  |  |  |  |  |  |
| Timiri | 59/624 (9.5%) | 2680/58662 (4.6%) | **2.18 [1.65, 2.83]\*\*\*** | 1.22 [0.91, 1.61] | Ref | Ref |
| Jawadhu Hills | 33/316 (10.4%) | 2853/22815 (12.5%) | 0.82 [0.56, 1.15] | 0.73 [0.48, 1.06] | **0.37 [0.24, 0.58]\*\*\*** | **0.50 [0.31, 0.81]\*\*** |
| **Pandemic** |  |  |  |  |  |  |
| year <= 2019 | 53/603 (8.8%) | 2176/51924 (4.2%) | **2.20 [1.64, 2.90]\*\*\*** | **1.64 [1.20, 2.18]\*\*\*** | Ref | Ref |
| year >= 2020 | 39/337 (11.6%) | 3357/29553 (11.4%) | 1.02 [0.72, 1.41] | **0.70 [0.48, 0.99]\*** | **0.46 [0.30, 0.72]\*\*\*** | **0.39 [0.25, 0.62]\*\*\*** |

**Table 3** presents the results of mixed-effects logistic regression analysis examining the associations between disability status and deworming treatment location among children who attended school and had received treatment. Overall, children with disabilities had significantly lower odds of receiving school-based treatment compared to children without disabilities (adjusted OR=0.57, 95% CI: 0.44-0.73) after adjusting for covariates. Subgroup analyses identified several significant interaction effects. Age emerged as a strong effect modifier (interaction adjusted OR=3.31, 95% CI: 1.99-5.59), indicating that older children (12-17 years) with disabilities had substantially higher odds of receiving school-based treatment compared to younger children with disabilities. Parental marital status demonstrated a significant interaction effect (interaction adjusted OR=0.36, 95% CI: 0.18-0.73), suggesting that children with disabilities whose parents were previously or never married had markedly lower odds of receiving school-based treatment compared to those with married parents. Distance to school also showed a significant interaction effect (interaction adjusted OR=0.45, 95% CI: 0.26-0.77), with children with disabilities living within 1km of school having lower odds of receiving school-based treatment compared to those living farther away.

**Table 3.** Association between disability status and treated in school, by overall and subgroups. A mixed-effects logistic regression analysis examining the association between disability status and deworming treated in school or community. The model fitted with treated in school (yes or no) as the dependent variable and disability status (yes or no) as the primary predictor, with adjustments for sociodemographic covariates including age, sex, household educational attainment, parental marital status, household size, socioeconomic status, caste, residential location, site, and survey round. Random intercepts were included for participant’s unique ID. Results are presented as odds ratios (OR) with corresponding 95% confidence intervals (CI). Analyses include overall population estimates and stratified subgroup analyses. Interaction effects between disability status and subgroup variables were assessed by including the interactive term between disability status and corresponding subgroup variable. n/N refers to the number of children treated in school (n) out of the total number of children who received deworming treatment in that category (N). For example, 230/319 indicates that 230 children with disability out of a total of 319 treated children with disability received their treatment in school.. p > 0.5, * p < 0.05, ** p < 0.01, *** p < 0.001

| Outcomes | Children with disability<br>n/N (%) | Children without disability<br>n/N (%) | Unadjusted OR | Adjusted OR | Interaction with subgroup |  |
| --- | --- | --- | --- | --- | --- | --- |
|  |  |  |  |  | Unadjusted OR | Adjusted OR |
| **Overall** | 230/319 (72.1%) | 36881/45245 (81.5%) | **0.59 [0.46, 0.75]\*\*\*** | **0.57 [0.44, 0.73]\*\*\*** | - | - |
| **By age** |  |  |  |  |  |  |
| 5-11 years | 82/137 (59.9%) | 16214/19580 (82.8%) | **0.31 [0.22, 0.44]\*\*\*** | **0.31 [0.22, 0.45]\*\*\*** | Ref | Ref |
| 12-17 years | 148/182 (81.3%) | 20667/25665 (80.5%) | 1.05 [0.73, 1.55] | 0.96 [0.67, 1.43] | **3.40 [2.06, 5.69]\*\*\*** | **3.31 [1.99, 5.59]\*\*\*** |
| **By gender** |  |  |  |  |  |  |
| Females | 121/163 (74.2%) | 18474/21969 (84.1%) | **0.55 [0.39, 0.78]\*\*\*** | **0.53 [0.37, 0.76]\*\*\*** | Ref | Ref |
| Males | 109/156 (69.9%) | 18407/23276 (79.1%) | **0.61 [0.44, 0.87]\*\*** | **0.60 [0.43, 0.86]\*\*** | 1.13 [0.69, 1.84] | 1.13 [0.68, 1.86] |
| **Highest education degree of family members** |  |  |  |  |  |  |
| No education | 71/103 (68.9%) | 7482/9116 (82.1%) | **0.48 [0.32, 0.75]\*\*\*** | **0.50 [0.33, 0.79]\*\*** | Ref | Ref |
| Any primary | 50/63 (79.4%) | 6920/8378 (82.6%) | 0.81 [0.45, 1.56] | 0.80 [0.44, 1.55] | 1.67 [0.81, 3.61] | 1.59 [0.76, 3.46] |
| Any middle | 41/55 (74.5%) | 8407/10062 (83.6%) | 0.58 [0.32, 1.10]. | 0.55 [0.30, 1.06]. | 1.19 [0.57, 2.55] | 1.06 [0.50, 2.29] |
| Any secondary or higher | 68/98 (69.4%) | 14072/17689 (79.6%) | **0.58 [0.38, 0.91]\*** | **0.55 [0.36, 0.87]\*\*** | 1.20 [0.66, 2.20] | 1.11 [0.60, 2.05] |
| **Parents marital status** |  |  |  |  |  |  |
| Married | 210/280 (75%) | 33612/41246 (81.5%) | **0.68 [0.52, 0.90]\*\*** | **0.66 [0.50, 0.87]\*\*** | Ref | Ref |
| Previously or never married | 20/39 (51.3%) | 3269/3999 (81.7%) | **0.24 [0.12, 0.45]\*\*\*** | **0.24 [0.12, 0.47]\*\*\*** | **0.34 [0.17, 0.69]\*\*** | **0.36 [0.18, 0.73]\*\*** |
| **Household socioeconomic status** |  |  |  |  |  |  |
| Rich (Upper 2 quintiles) | 138/185 (74.6%) | 23351/28952 (80.7%) | **0.70 [0.51, 0.99]\*** | **0.65 [0.47, 0.92]\*** | Ref | Ref |
| Poor (Lower 3 quintiles) | 92/134 (68.7%) | 13530/16293 (83%) | **0.45 [0.31, 0.65]\*\*\*** | **0.48 [0.33, 0.70]\*\*\*** | 0.64 [0.39, 1.04]. | 0.70 [0.42, 1.15] |
| **Distance to school** |  |  |  |  |  |  |
| <= 1km | 132/194 (68%) | 18422/22199 (83%) | **0.44 [0.32, 0.60]\*\*\*** | **0.42 [0.31, 0.57]\*\*\*** | **0.48 [0.28, 0.81]\*\*** | **0.45 [0.26, 0.77]\*\*** |
| >1km | 98/125 (78.4%) | 18459/23046 (80.1%) | 0.90 [0.60, 1.41] | 0.89 [0.58, 1.39] | Ref | Ref |
| **Residence place** |  |  |  |  |  |  |
| Rural | 209/281 (74.4%) | 31495/38323 (82.2%) | **0.63 [0.48, 0.83]\*\*\*** | **0.62 [0.48, 0.82]\*\*\*** | Ref | Ref |
| Urban | 21/38 (55.3%) | 5386/6922 (77.8%) | **0.35 [0.19, 0.68]\*\*\*** | **0.32 [0.17, 0.63]\*\*\*** | 0.56 [0.28, 1.13] | 0.53 [0.26, 1.08]. |
| **Site** |  |  |  |  |  |  |
| Timiri | 153/207 (73.9%) | 27809/34129 (81.5%) | **0.64 [0.47, 0.89]\*\*** | **0.61 [0.45, 0.84]\*\*** | Ref | Ref |
| Jawadhu Hills | 77/112 (68.8%) | 9072/11116 (81.6%) | **0.50 [0.33, 0.75]\*\*\*** | **0.53 [0.35, 0.81]\*\*** | 0.77 [0.46, 1.29] | 0.77 [0.46, 1.30] |

## Discussion

### Principle findings

This study highlights significant disparities in deworming treatment coverage between children with and without disabilities in the context of community-wide mass drug administration (cMDA) and school-based deworming. Overall, children with disabilities were less likely to receive deworming treatment compared to their typically developing peers, particularly during school-based deworming campaigns. While cMDA demonstrated relatively higher inclusivity by reaching children with disabilities outside the school system, treatment gaps persisted, underscoring systemic barriers to equitable healthcare access. These disparities were exacerbated by contextual factors such as age, parental marital status, distance to school, and geographic region, with older children, those from previously or never-married parents, and those residing in certain rural clusters experiencing the greatest inequities and the least likelihood of being dewormed. Additionally, during the COVID-19 pandemic, treatment patterns were substantially modified, with the impact of treatment access disruption being more pronounced among typically developing children compared to children with disabilities, revealing complex shifts in healthcare delivery during the pandemic period. These findings emphasize the need for targeted strategies to ensure equitable inclusion of children with disabilities in deworming campaigns, both in schools and the broader community.

### Compare with previous studies

Our findings support the concern that school-based health programs systematically exclude children with disabilities, thereby perpetuating health inequalities and potentially compromising the effectiveness of public health campaigns. School-based delivery platforms constitute essential mechanisms for providing preventive health interventions for children, including deworming, nutritional supplementation, immunization, and vision screening[19–21]. However, these approaches inherently disadvantage children with disabilities, who experience significantly lower rates of school enrollment and attendance compared to their non-disabled peers[22]. This systematic exclusion contributes to measurable health disparities, as demonstrated in studies where school-based health services represented primary access points for preventive care[23, 24]. For instance, research has documented how children with disabilities miss crucial nutritional interventions delivered through schools, subsequently exhibiting higher rates of malnutrition and associated health complications[25]. Multiple investigations across varied contexts have confirmed that children with disabilities face disproportionate barriers to healthcare access[26, 27], with school-based exclusion representing a significant contributing factor. The persistent marginalization of children with disabilities from school-based health interventions not only exacerbates existing health inequities, but may also undermine population-level public health initiatives by preventing the achievement of coverage thresholds necessary for intervention success.

Nevertheless, we found that gaps in treatment access for children with disabilities can be effectively addressed through community-based outreach efforts. Our findings demonstrate that community-wide mass drug administration approaches achieved higher inclusivity for children with disabilities compared to school-based deworming programs, challenging persistent assumptions that children with disabilities are inherently unreachable or hidden within communities. This observation aligns with emerging evidence suggesting that targeted community interventions can successfully mitigate the healthcare disparities experienced by marginalized populations[28]. While the primary objective remains expanding educational inclusion for children with disabilities—thereby facilitating access to both academic opportunities and associated health interventions—the reality necessitates complementary approaches. Integrating community outreach with school-based programs creates a comprehensive framework that ensures that health services reach children with disabilities, regardless of educational enrollment status. This dual-channel strategy not only promotes equitable health outcomes but also strengthens overall intervention effectiveness by capturing populations that would otherwise remain excluded. The successful implementation of community-wide deworming in our study demonstrates the feasibility of inclusive public health approaches that accommodate the needs of children with disabilities without requiring substantial infrastructure modifications or resource allocation.

The intersectional nature of disability and various social determinants substantially influences healthcare access patterns among children with disabilities. These children represent a remarkably diverse population across dimensions of age, gender, socioeconomic status, and geographic location, all of which are established determinants of health outcomes[29–31]. The significant site interaction effect (interaction adjusted OR=0.50, 95% CI: 0.31-0.81) observed in our study highlights how geographic factors, particularly distance to schools and challenging terrain in areas like Jawadhu Hills, disproportionately impact healthcare access for children with disabilities who may face mobility challenges. While the importance of examining intersectional factors in healthcare access is increasingly recognized, many studies remain underpowered to detect these complex interactions, resulting in limited evidence. Our research contributes significantly to this knowledge gap through robust assessment of effect modification across multiple dimensions. The findings reveal pronounced roles for social factors in mediating treatment access, including parental marital status and distance to school, alongside child-specific characteristics such as age. This pattern underscores the critical function of caregivers in facilitating healthcare access for children with disabilities, suggesting that effective interventions must extend beyond child-focused approaches to include caregiver support and education regarding preventive healthcare importance. The heterogeneity in treatment coverage observed across different subgroups challenges simplistic approaches to disability inclusion and advocates for nuanced strategies that address the multifaceted barriers experienced by diverse segments within this population[32–35].

The implications of our findings extend to both local and global contexts, necessitating targeted policy adjustments and programmatic innovations. For local policymakers and programme implementers in India, this research underscores the importance of redesigning the National Deworming Day initiative to incorporate robust community outreach components that specifically address children with disabilities who remain outside the educational system. Health workers and community drug distributors should receive specialized training on disability inclusion, with particular emphasis on identifying and reaching households with children with disabilities. Furthermore, coordination between education and health ministries must be strengthened to develop comprehensive databases that capture children with disabilities, regardless of school enrollment status. Globally, these findings hold significant relevance for other mass drug administration programs targeting various neglected tropical diseases in low- and middle-income countries where similar exclusion patterns are likely to exist. International organizations and funding bodies supporting such initiatives should incorporate disability-inclusive metrics into their monitoring frameworks and establish conditional funding mechanisms that incentivize inclusive programming. Additionally, the demonstrated effectiveness of community-wide approaches offers a valuable model for addressing health inequities beyond deworming—including vaccination campaigns, nutritional interventions, and other preventive services—potentially transforming how public health initiatives conceptualize and operationalize inclusivity for marginalized populations.

### Strength and limitations

This study has several strengths. It is one of the first studies to comprehensively evaluate disparities in deworming coverage among children with and without disabilities, leveraging high-quality, systematically collected data from the DeWorm3 trial, a large-scale, multi-country randomized study. The use of the Child Functioning Module (CFM), a validated tool grounded in the biopsychosocial model of disability, provides robust and standardized assessments of functional difficulty across multiple domains, ensuring the reliability of disability classification. Additionally, the inclusion of both community-wide mass drug administration (cMDA) and school-based deworming programs allows for a advanced comparison of these delivery platforms, shedding light on equity gaps in access to health interventions.

However, this study has limitations. First, the reliance on caregiver-reported data on disability through the CFM may have introduced reporting bias, particularly in communities where stigma around disability is prevalent. Moreover, the CFM uses a relatively high threshold of “a lot” of difficulty or more in at least one domain, and so may not capture all the children with disabilities. Second, while the study adjusted for a range of covariates, residual confounding may persist, especially for unmeasured factors, such as household attitudes toward disability or community-level barriers. Third, the generalizability of findings may be limited to settings similar to the DeWorm3 trial sites, particularly rural and tribal populations in Tamil Nadu, India. Finally, the impact of the COVID-19 pandemic on treatment coverage, while an important finding, complicates direct comparisons between school-based and community-based approaches, as the latter became the primary mode of treatment delivery during pandemic-related disruptions.

## Conclusion

School-based health programs may exclude children with disabilities, as evidenced by disparities in deworming coverage in southern India. While community-wide mass drug administration showed promise in reducing these inequities, gaps persisted, particularly for children with disabilities from specific demographic and geographic subgroups. The findings underscore the importance of combining school-based approaches with targeted community outreach to ensure comprehensive coverage. To achieve equitable health outcomes and successful STH transmission interruption, deworming programs must adopt disability-inclusive strategies. Future public health interventions should incorporate disability metrics in monitoring frameworks and design delivery approaches that accommodate children with diverse functional needs and living contexts.

## Contributors

SC and RMR contributed to the concept and study design. SC conducted the analysis. SC, RMR, KA, and SPK interpreted results. SC drafted the manuscript. SC, RMR, KA, SPK, and SA critically revised the manuscript for important intellectual content. All authors edited and approved the final manuscript.

## Data availability statement

Access to the Deworm3 data and supporting documents is available on request at vivli.org (https://doi.org/10.25934/PR00010754)

